# OUTCOME AND ASSOCIATED FACTORS AMONG ROAD TRAFFIC ACCIDENT VICTIMS AT WOLAITA SODDO CHRISTIAN HOSPITAL, WOLAITA SODDO, SOUTH ETHIOPIA 2020, INSTITUTION-BASED CROSS-SECTIONAL STUDY

**DOI:** 10.1101/2021.09.16.21262878

**Authors:** Asrat Hizkel, Teklemichael Gebru, Eshetu Yisihak, Desta Markos

## Abstract

**Background:** In today’ s world road traffic accident victims are treated as a major epidemic of non-communicable disease. Road traffic accidents caused numerous family tragedies such as serious economic loss to the community and the death of young people. The problem is more severe in low and middle-income countries. In Ethiopia, the largest proportion of series injuries comes from road traffic accidents and become major causes of death in the emergency room. Despite this, only a little is known about treatment outcomes of road traffic accident victims and its associated factors in Ethiopia.

**Method:** An institution-based Cross-sectional study design was conducted at Wolaita Soddo Christian hospital with a sample of 400 road traffic accidents. The medical record was selected using a systematic sampling method. Data was entered using Epi-data version 4.1 and was exported to and analyzed using SPSS version 23. A multivariable logistic regression model was used to assess the association between the independent variables and dependent variables.

**RESULT:** The overall death rate was 9.5%. Being out of hospital catchment area [AOR= 2.16, 95% CI= (1.01-4.70)] presence of co-morbid condition [AOR= 6.77 95% CI= (2.44-18.81)] lack of first aid help [AOR= 2.77 95% CI= (1.17-6.52)] and severity of the injury [AOR= 3.85 95% CI= (1.50-9.89)] were found to be significantly associated with outcome of road traffic accident victims.

**Conclusion:** The study shows that the death rate from road traffic accidents was high. Therefore, designing strategies to decrease death from road traffic accidents by giving great emphasis to road traffic accident victims with co-morbid conditions and severe injury and focusing on the availability and accessibility of pre-hospital care service.

## INTRODUCTION

Road traffic accident (RTA) is a collision between two or more vehicles, between vehicles and pedestrians, between vehicles and fixed obstacles (1). In today’ s world, RTA is treated as a major epidemic of non-communicable disease and incurs permanent disability, through amputation, head injury, or spinal cord injury. This causes numerous family tragedies and represents a serious economic loss to the community and the death of young people (2).

A road traffic accident is a growing public health problem, being responsible for up to 50 million nonfatal injuries globally (1). The burden of road traffic deaths is disproportionately high among low and middle than high-income countries concerning the size of their populations and the number of motor vehicles in circulation with an average rate of 27.5 deaths per 100,000 population, in low-income countries, and 8.3 deaths per 100,000 population in high -income countries(3). Ethiopia is one of the most affected countries by RTIs (4), with national RTAs fatality rate of 22% (5).

In Sub-Saharan Africa, there are challenges to delivering high-quality emergency medical services such as patient overload, poor integration with other health services, limited and inefficient services, poor clinical documentation, and a shortage of physicians and necessary supplies (6-8).

Even though Ethiopia is a country with a low rate of motorization, The number of people killed and injured as a result of traffic accidents has been increasing and the country was experiencing a tremendous loss of life and property each year as one of the leading countries of the world with worst accident record(9). In Ethiopia, the largest proportion of series injuries comes from road traffic accidents and become major causes of death in the emergency room among trauma victims (10).

Factors associated with outcomes of road traffic accidents were severity of the crash, time to admission, sex, time to take care, residence, age of the patient, admission systolic blood pressure <90 mmHg, severe head injury (Glasgow Coma Score = 3-8), time of receiving medical attention after injury, delayed presentation, had a comorbid condition, receiving first aid service (2, 11-17). In Ethiopia, outcomes and associated hospital factors were a little addressed area despite the increasing death secondary to RTAs. So, the aim of this study which assesses outcomes and associated factors among road traffic accident victims at Wolaita Soddo Christian hospital, south Ethiopia, attempts to fill this information gap.

## METHODS AND MATERIALS

### Study Area

Soddo town is the capital city of the Wolaita zone, which is one of 14 zones in the southern region of Ethiopia. Soddo town is 330 km far from Addis Ababa, the capital city of Ethiopia. Christian hospital is one of the two hospitals in Soddo town that gives trauma management at the highest level. The hospital has access to specialist and nursing care including emergency surgery, trauma surgery, critical care, neurosurgery, orthopedic surgery, anesthesiology, and radiology, as well as highly sophisticated surgical and diagnostic equipment.

### Study Period

May 10/2020 up to May 30/2020

### Study Design

Institution-based Cross-sectional study design

### Source population

All medical records of RTA victims admitted at Wolaita Soddo Christian hospital

### Study population

All medical records of RTA victims admitted at Wolaita Soddo Christian hospital from January 1, 2018, to December 31, 2019.

### Inclusion criteria

All RTA victims admitted from January 1, 2018, to December 31, 2019, at the emergency department of Wolaita Soddo Christian hospital were included in the study based on the sample size.

### Exclusion criteria

Those RTA victims’ deaths happened during arrival, immediate referral, incomplete cards (missing very important variable) were excluded from the study.

### Sample size determination

The sample size was determined by using single population proportion formula by considering the following assumptions: proportions of outcome from similar study 9.4%, 95% confidence level of certainty, 3% marginal error (we decreased the margin of error to increase sample size), and 10% non-response rate. The final sample size was 400.

### Sampling Technique

Using systematic random sampling, all RTA victims registered from January 1, 2018, to December 31, 2019, at Christian hospital were listed based on the sequence of their card numbers. Study units were selected by calculating sampling interval (K) from the sampling frame, N and sample size n (k=N/n), interval (k=5). The first number to start was selected randomly.

### Study Variables

#### Dependent variable

Outcome of road traffic accident / death /

#### Independent variable

Socio demographic characteristic, characteristics of RTA victims during admission to emergency department, Clinical characteristics at admission

#### Data collection instrument/ abstraction format/

The data abstraction format was developed in reference to the objective of the study. The data abstraction format was prepared in the English language.

#### Data collection process

The data for the study was review from routinely registered client medical records and emergency department registration books. The data was collected by trained clinical nurses under the supervision of investigators using a data abstraction format according to the inclusion criteria, the format contains questions regarding; socio-demographic, characteristics of RTA victims during admission to emergency department, clinical characteristics at admission, and treatment outcome.

#### Data Quality Control

To ensure the data quality, a pretest was conducted on 5% of the total sample. Reliabilities of the tool were checked. The data cleaning was done by running frequency and sorting by SPSS version 23.

#### Data Analysis

The collected data were entered into Epi data version 4.1 and analyzed using SPSS version 23. Frequency distribution, percentage calculation, means, and standard division were used to describe the variable. Those variables found to have a p-value <0.25 by binary logistic regression had been taken to multivariable logistic regression. Variables having p-values < 0.05 in the multiple logistic regression models were considered as significantly associated with the dependent variable.

## RESULT

### Socio-Demographic Characteristics

Of 400 RTA victims admitted to Soddo Christian hospital 232 (58%) were male.

The patients’ ages ranged from 1 to 85 years with the mean and standard deviations of 31.21 and ± 15 years respectively. About 308(77%) of RTA victims were from Wolaita while 92(23%) were out of Wolaita (Table 1).

**Table 1:**
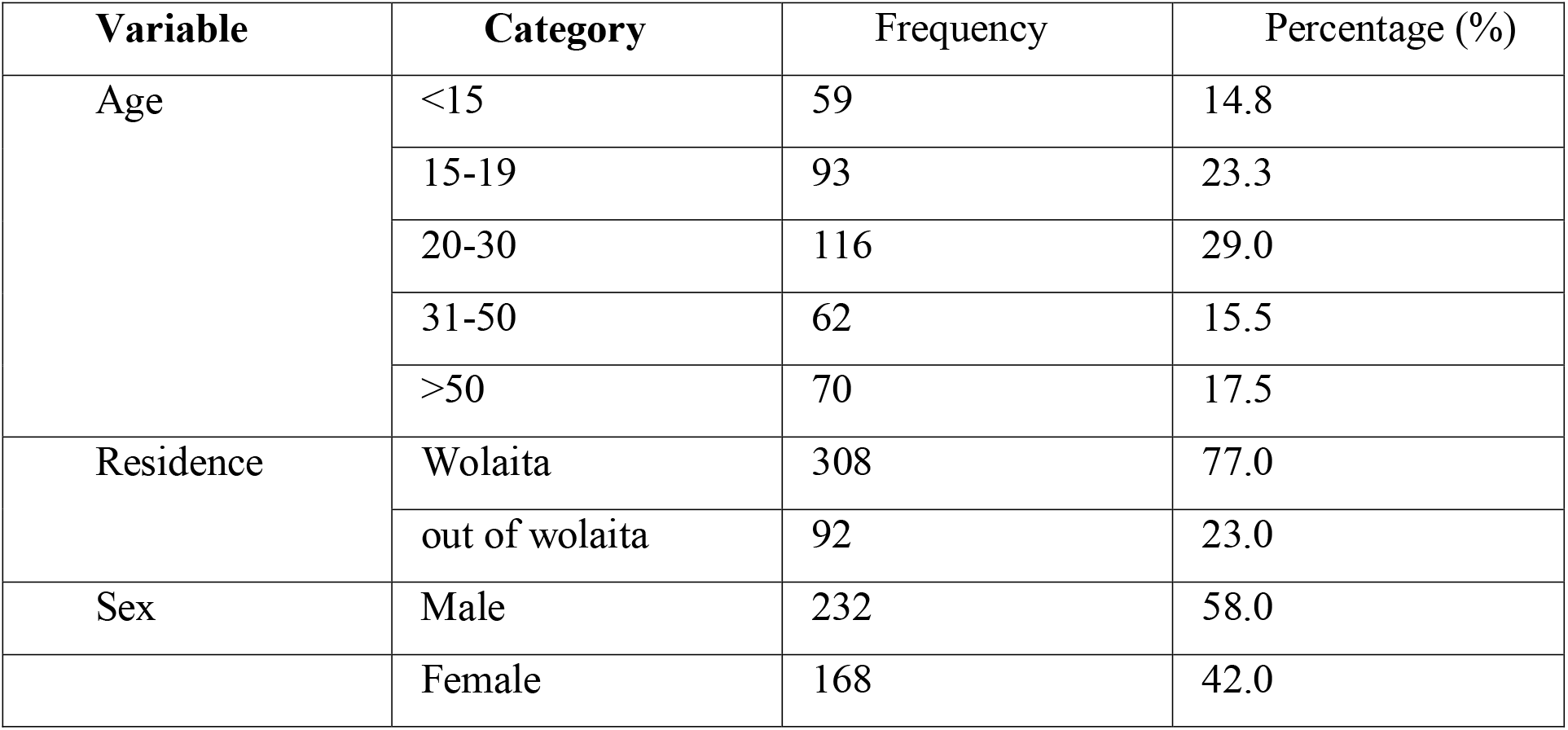
Distribution of demographic characteristics of RTA victims at Soddo Christian hospitals from January 1, 2018 -December 30, 2020

| Variable | Category | Frequency | Percentage (%) |
| --- | --- | --- | --- |
| Age | <15 | 59 | 14.8 |
|  | 15-19 | 93 | 23.3 |
|  | 20-30 | 116 | 29.0 |
|  | 31-50 | 62 | 15.5 |
|  | >50 | 70 | 17.5 |
| Residence | Wolaita | 308 | 77.0 |
|  | out of wolaita | 92 | 23.0 |
| Sex | Male | 232 | 58.0 |
|  | Female | 168 | 42.0 |

### Characteristics of RTA victims during admission to Emergency Department

About 149(37.3%) of the respondents come to the hospital within the first 6 hours of the accident while 38(9.5%) come to the hospital after 24 hours of the accident. More than half 240(60%) of the respondent arrived by ambulance while 10% of them arrived by carrying or walking. Only 160(40%) of the respondent had been given first aid before their arrival at Soddo Christian Hospital. About 202(50.5%) victims arrive at ED less than 24hr after injury and in 126(31.5%), 54 (13.5 %), 18 (9.5%) of the victims taken 1-3 days, 4-7 days, and beyond week respectively (Table 2).

**Table 2:** Characteristics of RTA victims during admission to the emergency department at Soddo Christian hospitals from January 1, 2018 -December 30, 2019

| Variable | Category | Frequency | Percentage (%) |
| --- | --- | --- | --- |
| Time of arrival to hospital after accident | 0hour-6hour | 149 | 37.3 |
|  | 6 hour - 12 hour | 102 | 25.5 |
|  | 12 hour - 18 hour | 73 | 18.3 |
|  | 18 hour -24 hour | 38 | 9.5 |
|  | after 24 hour | 38 | 9.5 |
| Mode arrival to hospital | Ambulance | 240 | 60.0 |
|  | Taxi | 120 | 30.0 |
|  | Other /carrying, walking/ | 40 | 10.0 |
| First aid given | Yes | 160 | 40.0 |
|  | No | 240 | 60.0 |
| Time gap between accident and arrival | <24 hour | 202 | 50.5 |
|  | 1-3 days | 126 | 31.5 |
|  | 4-7 days | 54 | 13.5 |
|  | > 1 week | 18 | 4.5 |
| Source of referrals | Governmental hospitals in Wolaita Soddo |  | 31.75 |
|  | Governmental hospitals out of Wolaita Soddo |  | 10.5 |
|  | Private hospitals/clinics/ in Wolaita Soddo |  | 21 |
|  | Private hospitals/clinics/ out of Wolaita Soddo |  | 10.5 |
|  | Health center |  | 15.75 |
|  | Self-referral |  | 10.5 |

### Clinical characteristics of RTA victims at admission

Among the respondents admitted to Soddo Christian frequently hospital injured body regions were lower extremity 75(18.5%) and abdominal area 69(17.3%). For the remaining RTA Victims, the injury involves the upper extremity, chest, pelvic, head, and other: 67 (16.8%), 44 (11%), 50 (12.5%), 62 (15.5%), and 33 (8.3%) of the victims correspondingly. When we look at the level of severity 203 (50.8%) were mild, 82 (20.5%) were moderate, 54 (13.5%) severe pain levels were had and 61 (15.3 %) victims were classified as needing immediate intervention. Were as 160 (40%) RTA victims were receiving first-aid and 240 (60%) RTA victims did not receive the service in the present study, in addition, 35(8.8%) of RTA, Victims admitted to ED had co-morbid conditions (Table 3).

**Table 3:** Clinical characteristics of RTA victims during admission at Soddo Christian hospital from January 1, 2018 -December 30, 2019

| Variable | Category | frequency | Percentage (%) |
| --- | --- | --- | --- |
| have co morbid condition | Yes | 35 | 8.8 |
|  | No | 365 | 91.3 |
| First aid given | Yes | 160 | 40.0 |
|  | No | 240 | 60.0 |
| Level of severity | Green color coding | 203 | 50.8 |
|  | Yellow color coding | 82 | 20.5 |
|  | Orange color coding | 54 | 13.5 |
|  | Red color coding | 61 | 15.3 |
| Site of injury | Head injury | 62 | 15.5 |
|  | Upper extremity | 67 | 16.8 |
|  | Chest injury | 44 | 11.0 |
|  | Abdominal injury | 69 | 17.3 |
|  | Pelvic injury | 50 | 12.5 |
|  | Lower extremity injury | 75 | 18.8 |
|  | Other /multiple organ injury ,spinal cord injury/ | 33 | 8.3 |

### Outcomes of road traffic accident victims at Christian hospital

Of the total RTA victims, about 362 (90.5 %) were discharged with medical advice, and 38 (9.5%) died during the course of the treatment (Figure1).

### Factors associated with RTA related death

Road traffic accident victims those lives out of Wolaita (far), were 2.16 times more likely to die, (AOR=2.16: 95% CI: 1.01-4.70) than RTA victims who came to the Soddo Christian hospital from the Wolaita area(near). RTA victims those, had co-morbid conditions, were 6.77 times more likely to die than RTA victims who had not co-morbid conditions (AOR: 6.77; 95% CI (2.44-18.81). According to the study, RTA victims who had not received first aid was 3 times more likely to die than the respondent who had received first aid (AOR: 2.77; 95% CI (1.17-6.52). RTA victims who had been severely injured were more likely to die than those who had been affected slightly (AOR: 3.85; 95% CI (1.50-9.89) (Table 4).

**Table 4:**
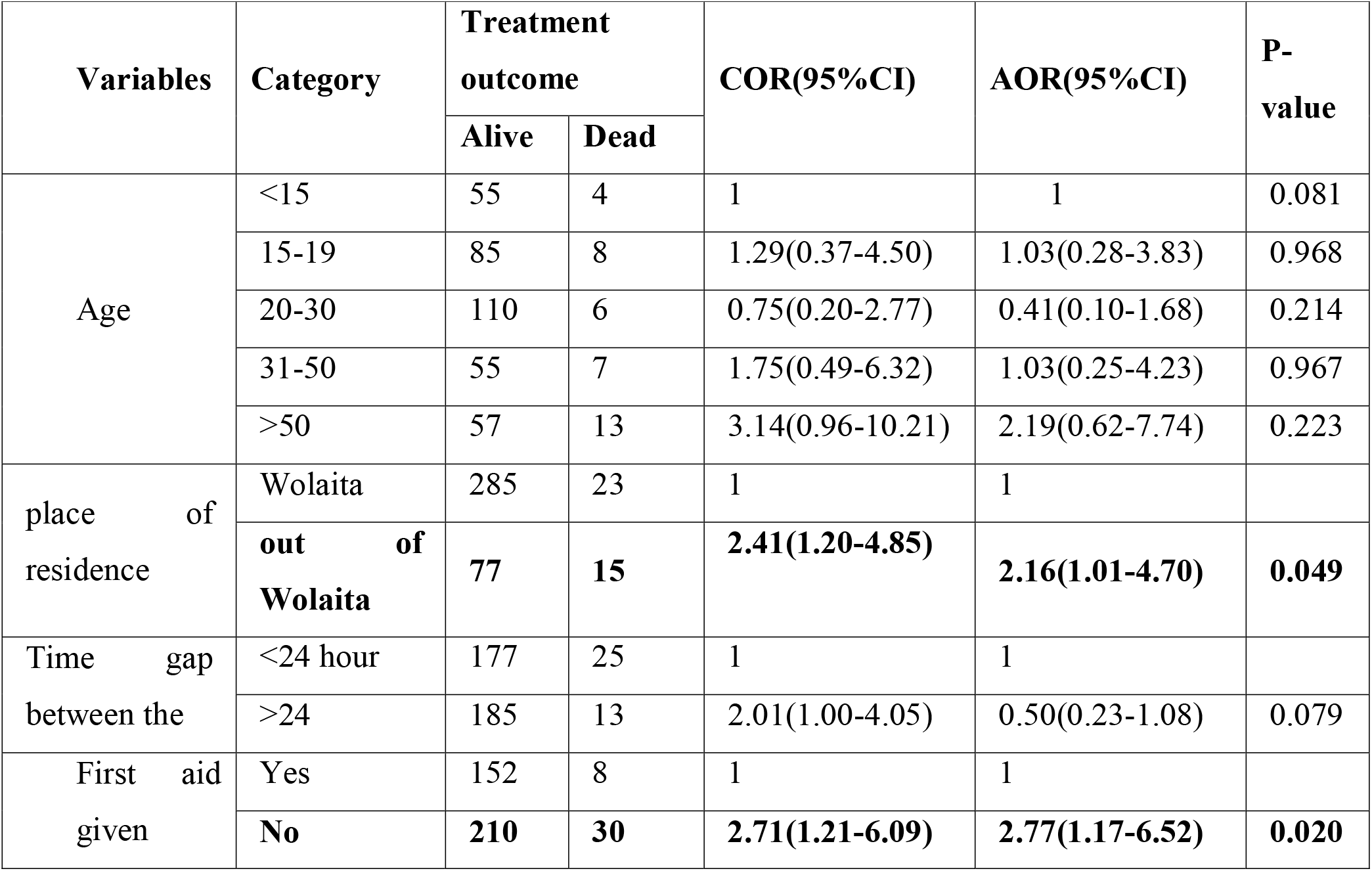
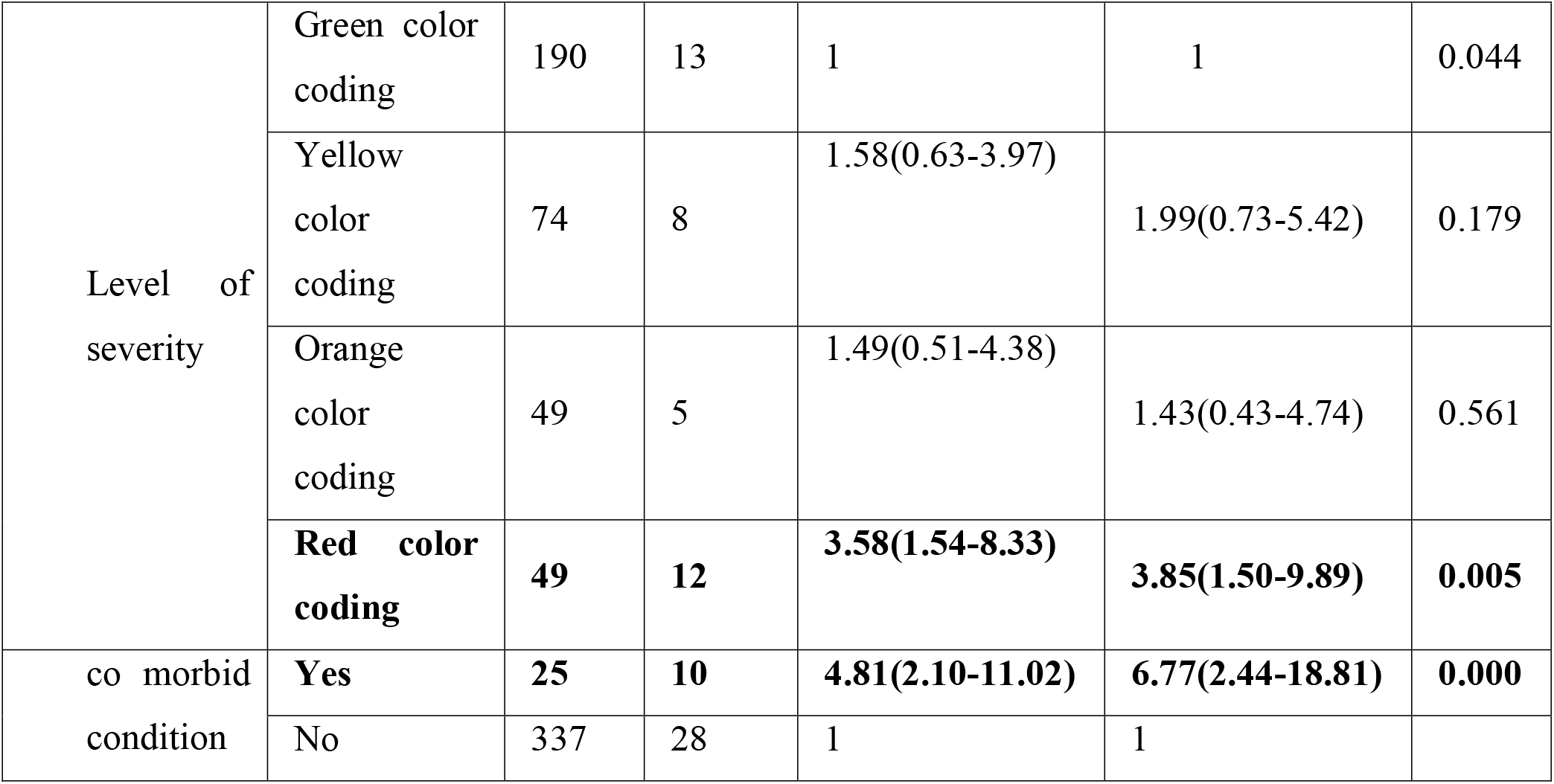
Factors associated with death among RTA victims during admission at Soddo Christian hospitals from January 1, 2018 -December 30, 2019

| Variables | Category | Treatment outcome |  | COR(95%CI) | AOR(95%CI) | P-value |
| --- | --- | --- | --- | --- | --- | --- |
|  |  | Alive | Dead |  |  |  |
| Age | <15 | 55 | 4 | 1 | 1 | 0.081 |
|  | 15-19 | 85 | 8 | 1.29(0.37-4.50) | 1.03(0.28-3.83) | 0.968 |
|  | 20-30 | 110 | 6 | 0.75(0.20-2.77) | 0.41(0.10-1.68) | 0.214 |
|  | 31-50 | 55 | 7 | 1.75(0.49-6.32) | 1.03(0.25-4.23) | 0.967 |
|  | >50 | 57 | 13 | 3.14(0.96-10.21) | 2.19(0.62-7.74) | 0.223 |
| place of residence | Wolaita | 285 | 23 | 1 | 1 |  |
|  | out of Wolaita | 77 | 15 | 2.41(1.20-4.85) | 2.16(1.01-4.70) | 0.049 |
| Time gap between the | <24 hour | 177 | 25 | 1 | 1 |  |
|  | >24 | 185 | 13 | 2.01(1.00-4.05) | 0.50(0.23-1.08) | 0.079 |
| First aid given | Yes | 152 | 8 | 1 | 1 |  |
|  | No | 210 | 30 | 2.71(1.21-6.09) | 2.77(1.17-6.52) | 0.020 |
| Level of severity | Green color coding | 190 | 13 | 1 | 1 | 0.044 |
|  | Yellow color coding | 74 | 8 | 1.58(0.63-3.97) | 1.99(0.73-5.42) | 0.179 |
|  | Orange color coding | 49 | 5 | 1.49(0.51-4.38) | 1.43(0.43-4.74) | 0.561 |
|  | **Red color coding** | **49** | **12** | **3.58(1.54-8.33)** | **3.85(1.50-9.89)** | **0.005** |
| co morbid condition | **Yes** | **25** | **10** | **4.81(2.10-11.02)** | **6.77(2.44-18.81)** | **0.000** |
|  | No | 337 | 28 | 1 | 1 |  |

## Discussion

The study revealed that the outcome of 9.5% of RTA victims admitted to Soddo Christian hospitals during the study period was death. The finding of this study is in line with a study from Dilchora referral hospital, Dire Dawa (9.4%) (18).

The death outcome observed in this study was higher as compared to the studies in hospitals in Ahmedabad city, India (1.33%) (19) and (3.9%) in Iran (20), Zewuditu memorial hospital (1%) (21), and Wolaita Zone, SNNPR 6 % (22). This might be due to the studies set up was trauma treatment centers and many victims were linked from another health facility, due to this patient overload and life-treating cases may lead to higher deaths.

However, it is lower than a report from a previous study conducted (13.1%) at the tertiary care center, India (23). It might be due that Soddo Christian hospital mainly works on orthopedic surgery in that trauma management was given at the highest levels in the study area.

The study result showed that there was no significant association between sex, age, source of referral, mode of arrival, time of arrival, and site of injury with outcome /death / of RTA victims in Soddo Christian hospital but there was a significant association between residence, presence of first aid, presence of the co-morbid condition, and severity of the injury and treatment outcome /death / of RTA victims in Soddo Christian hospital.

Road traffic accident victims who live out of Wolaita were more likely to die than RTA victims who came to the hospital from the Wolaita area. This result is in line with the studies done in North Ghana, the death rate was 2 times increased from those coming from a rural area or out of hospital catchment (11, 12). This might be justifying the fact that when the victim comes far from the hospital time to admission was higher in comparison with those who come from near to hospital or treatment center this in turn increases the severity of the injury and poor outcome of RTA victims.

RTA victims those, had co-morbid conditions, were more likely to die than RTA victims who had no co-morbid conditions. The previous study was done in a tertiary health care delivery institute in western Nepal (24) also supports the finding as co-morbid conditions were significantly associated with a high death rate. Co-morbid conditions by themselves suppress the immunity and hinder recovery might be lead to death.

According to the study, RTA victims who had not received first aid were more likely to die. It is supported by a study conducted in tertiary care center Luck now, India, it indicates the proportion of death was significantly higher in those patients who got first aid in 30 minutes to 60 minutes as compared to those patients who got it in less than 30 minutes (19). This might be justified, not reserving first aid were decreased the chance of survival by complicated the case.

RTA victims who were at a red color-coding level of severity (severe pain level and victims with immediate intervention need injury) were more likely to die. The finding was similar to the study done in Iran; those RTA victims severely injured were more likely to die (20). A 5-year retrospective study done in India, also stated that the severity of injuries causes road traffic death (19). This might be justifying because physiological, anatomical, and psychological trauma caused by RTA leads to an unsatisfactory response for treatment modality.

## Conclusion

The study revealed among a hundred RTA victims’ approximately ten deaths was happen. Co-morbid condition, residence, the severity of the injury, not received first aid service were statistically significant factors that increase the death of RTA victims. It is better if the health care providers emphasize those RTA victims with co-morbid conditions and severe injury and Wolaita zone health bureau and federal ministry of health focusing on the availability and accessibility of pre-hospital care service.

## Data Availability

All data that support these findings are readily available.

## ABBREVIATIONS AND ACRONYMS

CI: Confidence Interval
AOR: Adjusted Odd Ratio
ED: Emergency Department
ICU: Intensive Care Unit
MEWS: Modified Early Warning Score
RTA: Road Traffic Accident
RTC: Road Traffic Crash
RTI: Road Traffic injury
SPSS: Statistical Package for Social Science

## Availability of data and materials

The dataset supporting the conclusions of this article is included within the article.

## ACKNOWLEDMENTS

The authors thank Wolaita Soddo Christian hospital and Wolaita zone health bureau for their cooperation by giving us important information, as well as data collectors.

## Authors’ contributions

**Conceptualization:** Asrat Hiskel, Eshetu Yisihak, Desta Maskos, Teklemichael Gebru

**Data curation:** Asrat Hiskel, Eshetu Yisihak, Desta Maskos, Teklemichael Gebru

**Formal analysis:** Asrat Hiskel, Eshetu Yisihak, Desta Maskos, Teklemichael Gebru

**Funding acquisition:** Asrat Hiskel, Eshetu Yisihak, Desta Maskos, Teklemichael Gebru

**Methodology:** Asrat Hiskel, Eshetu Yisihak, Desta Maskos, Teklemichael Gebru

**Project administration:** Asrat Hiskel, Eshetu Yisihak, Desta Maskos, Teklemichael Gebru

**Resources:** Asrat Hiskel, Eshetu Yisihak, Desta Maskos, Teklemichael Gebru

**Software:** Asrat Hiskel, Eshetu Yisihak, Desta Maskos, Teklemichael Gebru

